# Study protocol for a pilot clinical trial to understand neural mechanisms of response to a psychological treatment for pain and anxiety in pediatric functional abdominal pain disorders (FAPD)

**DOI:** 10.1101/2024.02.08.24302498

**Authors:** Natoshia R. Cunningham, Michelle A. Adler, Brittany N. Barber Garcia, Taylor Abounader, Alaina K. Miller, Mariela Monzalvo, Ismaeel Hashemi, Ryan Cox, Samantha L. Ely, Yong Zhou, Mark DeLano, Todd Mulderink, Mathew J. Reeves, James L. Peugh, Susmita Kashikar-Zuck, Robert C. Coghill, Judith E. Arnetz, David C. Zhu

**Author notes:** All authors had a substantive role in manuscript completion.

## Abstract

**Background:** Functional abdominal pain disorders (FAPD) are the most common chronic pain conditions of childhood and are made worse by co-occurring anxiety. Our research team found that the Aim to Decrease Pain and Anxiety Treatment (ADAPT), a six-session coping skills program using cognitive behavioral therapy strategies, was effective in improving pain-related symptoms and anxiety symptoms compared to standard care. In follow-up, this current randomized clinical trial (RCT) aims to test potential neural mechanisms underlying the effect of ADAPT. Specifically, this two-arm RCT will explore changes in amygdalar functional connectivity (primary outcome) following the ADAPT protocol during the water loading symptom provocation task (WL-SPT). Secondary (e.g., changes in regional cerebral blood flow via pulsed arterial spin labeling MRI) and exploratory (e.g., the association between the changes in functional connectivity and clinical symptoms) outcomes will also be investigated.

**Methods:** We will include patients ages 11 to 16 years presenting to outpatient pediatric gastroenterology care at a midwestern children’s hospital with a diagnosis of FAPD plus evidence of clinical anxiety based on a validated screening tool (the Generalized Anxiety Disorder-7 [GAD-7] measure). Eligible participants will undergo baseline neuroimaging involving the WL-SPT, and assessment of self-reported pain, anxiety, and additional symptoms, prior to being randomized to a six-week remotely delivered ADAPT program plus standard medical care or standard medical care alone (waitlist). Thereafter, subjects will complete a post assessment neuroimaging visit similar in nature to their first visit.

**Conclusions:** This small scale RCT aims to increase understanding of potential neural mechanisms of response to ADAPT. ClinicalTrials.gov registration: NCT03518216

## Introduction

### 1.1. Background

Functional abdominal pain disorders (FAPD) are among the most common chronic pain conditions impacting children and adolescents. These complex pain conditions are thought to result from dysregulations in the brain-gut axis and are characterized by aberrant amygdalar functional connectivity patterns when experiencing pain during a water loading symptom provocation task (WL-SPT), and at rest when compared to healthy youth [1]. The prevalence of such conditions ranges from 4% to 41% globally and the median rate is approximately 12% [2]. These conditions account for 50% of all pediatric gastroenterology (GI) visits [3]. One third of youth with FAPD experience impairment over the long term (e.g., for five years or more) [4], and impairment may be more pronounced in adolescent versus pre-adolescent youth [5]. Anxiety is also very common in youth with FAPD, with prevalence between 42% to 85% [5–7]. Youth with FAPD and comorbid anxiety have greater rates of pain-related impairment [8] and higher pain intensity [5]. Those who have elevated levels of anxiety, pain and disability are at risk for persistently elevated disability [8]. Further, youth with chronic pain conditions such as FAPD and comorbid anxiety are less likely to respond to cognitive behavioral therapy for pain management, the current gold standard for care [9].

To improve outcomes for youth impacted by FAPD and comorbid anxiety, the study PI and her team developed an enhanced approach to care, called the Aim to Decrease Anxiety and Pain Treatment (ADAPT) [10]. ADAPT is a six-session, individually delivered protocol that targets pain and anxiety through a blend of live and web-based sessions drawn from cognitive behavioral therapy [11, 12] and in the current study have also been infused with mindfulness meditation approaches [13]. ADAPT was found to be superior in reducing symptoms of functional disability and anxiety in youth with FAPD in a randomized clinical trial as compared to standard medical treatment alone [14]. Although the study team has previously found evidence of aberrant functional connectivity in the brains of youth with FAPD [1] (during the WL-SPT and when compared to the brains of healthy youth), the potential neural correlates underlying effective *treatments* for these conditions are unknown. This is important because it allows us to understand whether an effective nonpharmacologic treatment for pain and anxiety impacts neurological functioning in youth with FAPD. Here we report the protocol for a randomized trial designed to investigate the potential neural mechanisms associated with the treatment effect of ADAPT.

### 1.2. Study aims and hypotheses

The goal of this randomized controlled trial (RCT) is to investigate potential neural mechanisms underlying the treatment effect of ADAPT. Specifically, it is hypothesized that ADAPT will correspond to the normalization of hyperactive functional connectivity patterns (following at WL-SPT and at rest) between the amygdala and other regions implicated in pain as compared to those undergoing standard care/waitlist control condition. Secondarily, we propose that regional cerebral blood flow (CBF) observed via 3D Pulsed Arterial Spin Labeling (PASL) MRI will be reduced following ADAPT versus standard care/waitlist (after the WL-SPT and at rest). Lastly, reductions in functional connectivity and regional CBF will correspond to reductions in child-reported pain intensity and unpleasantness symptoms. This paper details the study protocol of this clinical trial, NCT03518216, funded by a National Center for Complementary and Integrative Health/National Institutes of Health (NCCIH/NIH) K23 grant, AT009458.

#### Primary Outcomes

*Aim 1a*. Left amygdala-prefrontal cortex (AMY-PFC) functional connectivity will be significantly reduced (i.e., evidence of decreased hyperconnectivity) after the WL-SPT post ADAPT vs. waitlist. *Aim 1b*. Changes between both the left and right amygdala and other network regions (e.g., thalamus) that are implicated in pain processing (e.g., default mode network [1, 15–18]) following ADAPT versus waitlist will also be investigated via a whole brain search. This includes changes following the WL-SPT and differences during rest between the ADAPT and waitlist groups.

#### Secondary Outcomes (Aim 2)

To assess if regional CBF reduction is associated with cognitive (e.g., prefrontal cortex [PFC]), affective (e.g., pregenual anterior cingulate cortex [pgACC], amygdala [AMY]), and visceral afferent (e.g., insula [INS], thalamus, anterior mid-cingulate cortex [aMCC], S1 & S2) pain. It is hypothesized that regional CBF associated with cognitive, affective, and visceral afferent pain will be significantly reduced (following the Wl-SPT and at rest) after ADAPT vs. waitlist.

#### Exploratory Outcomes

To assess if changes in functional connectivity and brain activations following ADAPT will correspond to reductions in pain (intensity and unpleasantness) and anxiety ratings. Our previous research has indicated amygdalar functional connectivity changes in youth with FAPD undergoing water loading are evident when accounting for changes in pain intensity and pain unpleasantness [1].

As noted below, additional measures of psychosocial factors (e.g., anxiety, pain catastrophizing, adverse childhood experiences), physical impairment (e.g., functional disability, pain interference) and sociodemographic data will also be collected.

## Methods

### 2.1. Study design and setting

The current study is a masked and mechanistic pilot clinical trial. The primary study site (Michigan State University [MSU], led by NRC) will coordinate the study activities. Any modifications will be communicated with IRB, study team, data monitoring committee, and the NCCIH as appropriate.

Study recruitment of male and female youth will take place in outpatient pediatric gastroenterology clinics at Helen DeVos Children’s Hospital/ (HDVCH)/Corewell Health. In-person and virtual recruitment will be utilized depending on whether the patient presents to clinic in person or virtually (see section *2.3* *Study Procedures* for more details). Patients diagnosed with FAPD by the medical team who agree to participate will complete additional screening measures to assess subject eligibility (see section *2.2**. Eligibility criteria* for more details).

Qualifying participants will complete two neuroimaging assessments with surveys (baseline and post, approximately 8 weeks part) (see Fig 1) at an outpatient imaging clinic at a satellite location of Corewell Health where the study scanner is located (approximately 15 minutes from the main children’s hospital).

**Fig 1.**
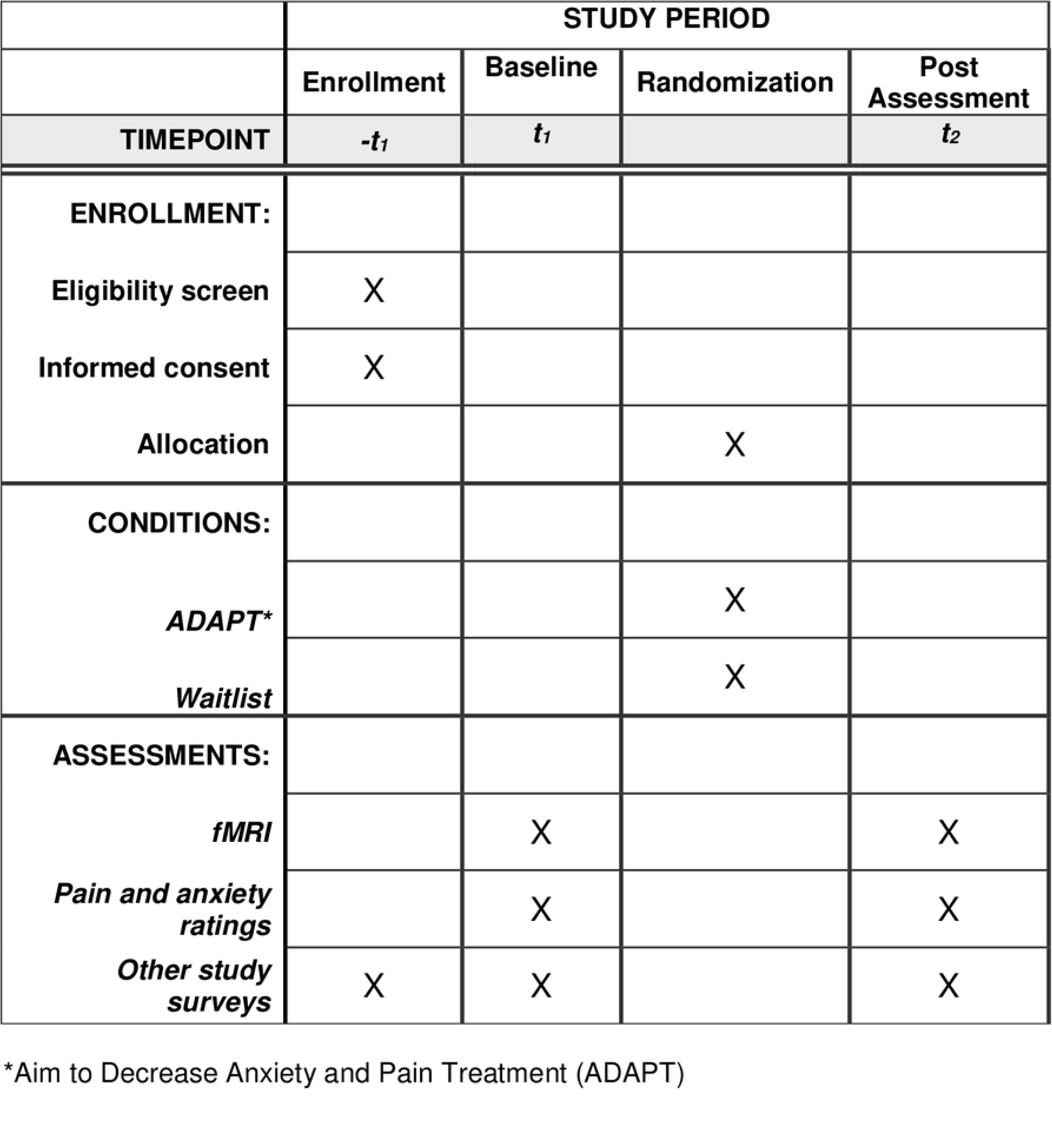
Schedule of Enrollment, Interventions, and Assessments. SPIRIT schedule of participation.

Following the baseline assessment, participants will be randomized to ADAPT or waitlist using a concealment method by a staff external to the study team (see section *2.4* *Randomization)*. The outcome assessor, study PI and study statisticians will be blinded to participant randomization. The blind will be broken at the clinical discretion of the supervising psychologist (BBG). Circumstances for breaking the blind may include cases when a patient’s safety is at risk, differential attrition, or if a patient’s symptoms remarkably worsen throughout treatment. If the blind needed to be broken, the study team would communicate with the local IRB, any NCCIH members involved in the study, and involve our study team, included but not limited to the PI, co-mentors, the biostatistician, and the outcome accessors (post-baccalaureate research coordinator).

The ADAPT program (see section *2.6* *Intervention* for more details) will be delivered virtually by a trained psychological provider (advanced doctoral students of pediatric psychology (TA, AM, MM)) under the supervision of a licensed clinical pediatric pain psychologist (BBG). Of note, live portions of the original ADAPT protocol were originally delivered in-person, however, due to COVID-19, we will deliver those portions of ADAPT using HIPAA compliant Zoom videoconferencing.

The target population is male and female youth between the ages of 11-16 with functional abdominal pain disorders (FAPD) and clinically significant anxiety based on the Generalized Anxiety Disorder-7 [GAD-7] [19] measure with a total cut-off score ≥ 10; (see section *2.5**. Assessments & Outcome Measures* for additional details). We aim to have between 17–25 participants in each arm for a total N of 34-50 participants in the study. Gender and age will be used as blocking variables in randomization (see more details in section *2.4* *Randomization*).

We plan to approach approximately 124 participants, ages 11-16 years. Of those, we expect 75% will agree to participate (n=93) based on our pilot functional magnetic resonance imaging (fMRI) study [10]. Of those, we expect approximately 65% will qualify. Thus, we anticipate recruiting n=60 to complete a baseline assessment. Based on our previous RCT, we expect 85% of those recruited will complete the baseline assessment (n = 50) and the majority will be retained in the study (90%, n=45). Based on our fMRI pilot study, we expect to lose approximately 10% of participant data to movement artifacts yielding n=40 with usable data.

This study is approved by Michigan State University’s institutional review board, (IRB), STUDY00003419, with Helen DeVos Children’s Hospital/Corewell Health relying on the primary IRB.

### 2.2. Eligibility criteria

*Inclusion criteria* will include children who must: 1) be between 11 and 16 years of age and their parent/primary caregiver will participate in the study; 2) meet criteria for FAPD based on physician/nurse practitioner diagnosis of FAPD and ROME IV FAPD criteria [20, 21] (see section *2.5**. Assessments & Outcome Measures* for additional details); 3) meet criteria for presence of clinical anxiety (based on the Generalized Anxiety Disorder-7 [GAD-7] [19] measure with a total cut-off score ≥ 10; (see section *2.5**. Assessments & Outcome Measures* for additional details)) or a current diagnosis of an anxiety disorder as indicated in the patient electronic medical record; 4) have sufficient English language ability (along with their caregiver) necessary to complete the study measures and protocol.

*Exclusion criteria* will include patients with: 1) a significant medical condition(s) with an identifiable organic cause including those that may include abdominal pain symptoms (e.g., Inflammatory Bowel Diseases); 2) a documented developmental delay(s), autism spectrum disorder, a previously diagnosed thought disorder (i.e., psychosis), or bipolar disorder; 3) significant visual, hearing, or speech impairment; organic brain injury; 4) current psychological therapy for pain or anxiety; 5) depressive symptoms cut-off score ≥ 22 on the Patient Health Questionnaire-9 [PHQ-9] [22] (see section *2.5**. Assessments & Outcome Measures* for additional details), or current active suicidal ideation; 6) an implant such as a cochlear implant device, a pacemaker or neurostimulator containing electrical circuitry or generating magnetic signals. Participants with 7) any significant ferrous material in their body that could pose the potential for harm in the MRI environment or cause signal dropout in key brain regions (i.e. orthodontia); 8) reported current/suspected pregnancy; 9) evidence of claustrophobia; 10) inability or unwillingness of individual or legal guardian/representative to give written informed consent.

### 2.3. Study procedures

Eligible participants with FAPD will be identified for the study from new or existing patients seen at the outpatient pediatric GI clinics. The lead GI care provider (e.g., physician, nurse practitioner) will confirm that identified patients meet criteria for FAPD, introduce the study, and then study staff will speak with the family to provide additional details and obtain verbal and written consent (caregiver) and assent (youth participant). If study staff are not physically present (i.e., if the GI visit occurs via telehealth, or if study staff are unable to attend the GI visit), virtual consent procedures will be conducted via phone and Research Electronic Data Capture (REDCap) eConsent Framework to minimize patient risk (includes written consent). Written consent will be obtained from the participating caregiver, and written assent will be obtained from the participating child. After consent is obtained, study staff will conduct a psychosocial screening to determine eligibility if screening was not performed as part of the clinic visit (e.g., anxiety and depressive symptom screening); (see section *2.5**. Assessments & Outcome Measures* and Fig 2 for additional details). Of note, the HDVCH/Corewell healthcare system routinely screens patients for anxiety and depression using the PHQ-4 [23] as part of standard medical care, with the GAD-7 [19] and PHQ-9 [22] subsequently administered for those who endorse anxiety or depressive symptomatology. Even if children do not endorse symptoms on the PHQ-4, the research team will administer the GAD-7 and PHQ-9 to interested and potentially eligible participants following the consenting process.

**Fig 2.**
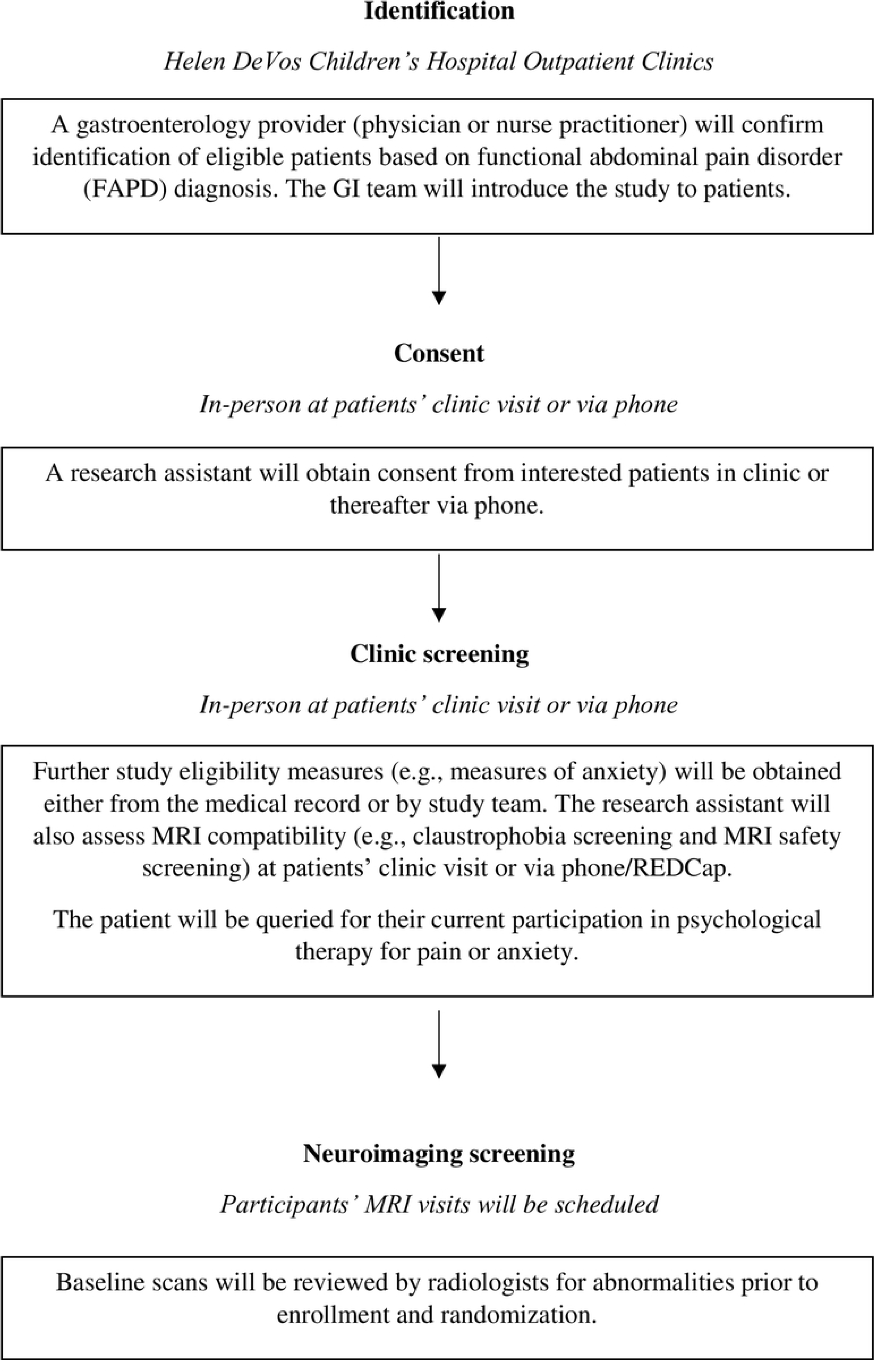
Referral, screening, and enrollment procedure. The medical provider will confirm patient diagnosis of FAPD and introduce the study. If a patient is interested, the research assistant will follow-up with the patient and caregiver (in-person at the clinic visit or via phone). The research assistant will perform consent procedures and complete screening measures that were not a part of the patients’ medical visit. After eligibility screening, the patient will be enrolled.

Potentially eligible participants will also be screened for MRI eligibility (e.g., to rule out claustrophobia and presence of metal in the body). Enrolled patients will then be scheduled for their MRI visits, approximately 8 weeks apart (see Fig 3). For participating, youth will receive up to $200 in Amazon gift cards ($100 at baseline and $100 at the post assessment). If a participant elects not to complete ADAPT, they are still eligible to complete the post-assessment.

**Fig 3.**
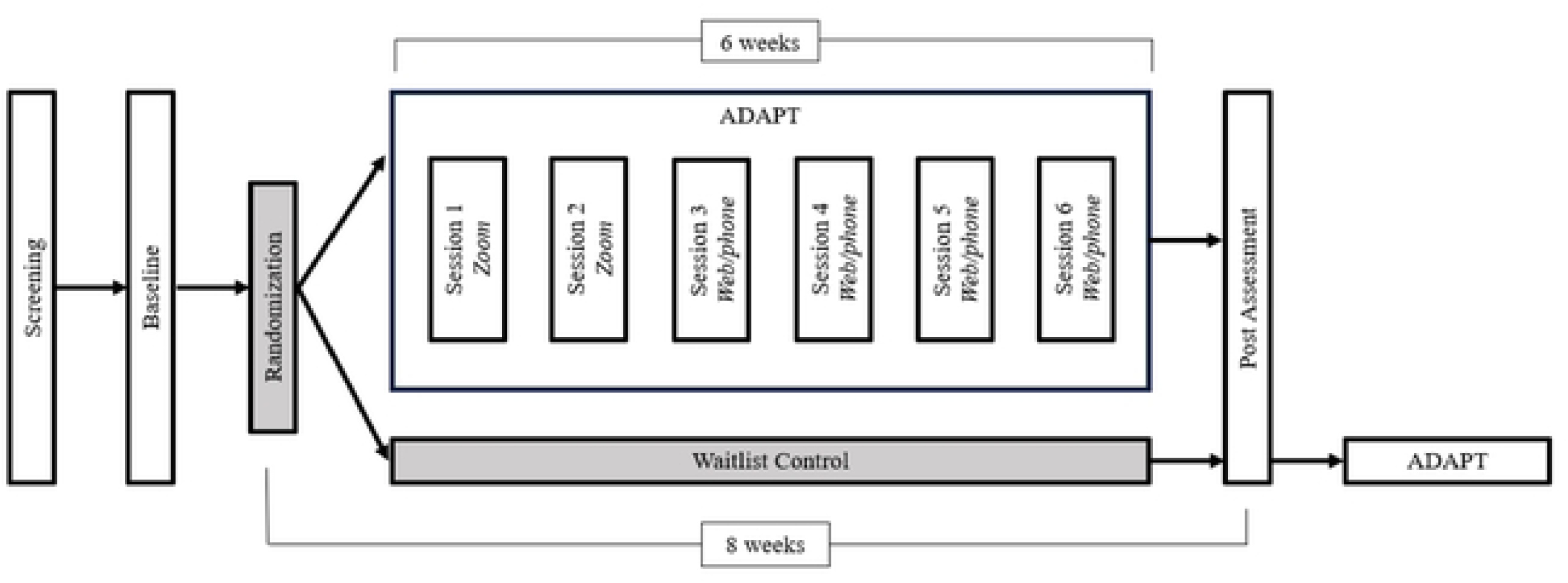
Study timeline. Participants will be screened and if eligible, will be scheduled for a baseline assessment. Following the baseline visit, participants will be randomized to either ADAPT or a waitlist control group. Participants will complete a post assessment ∼7-9 weeks from the baseline assessment. Participants who are part of the waitlist control group will be offered ADAPT following their post assessment as a clinical courtesy, which is detailed in the consent process.

### 2.4. Randomization

Following the baseline visit, MRI images will be reviewed by a radiologist (TM, MD) for safety. Assuming no incidental findings which would preclude participation are present, participants will be randomized. Randomization will be generated using PROC PLAN in SAS 9.3 by an external team member. Gender and age will be used as blocking variables in randomization. The randomizer will not have access to the data. The randomization schedule and code sheet will be held confidentially by a designated secondary research team member not involved in data collection, and the study interventionist will contact the designee to get the next assignment when a participant is enrolled in the study. The outcome assessor, study PI, and study statistician will be blinded to participant randomization. Patients and their families will be informed of group assignment within one week of their baseline assessment and begin ADAPT or waitlist approximately one week after group assignment.

### 2.5. Assessments & Outcome Measures

As noted above, qualifying participants and their respective caregivers will be invited to attend an in-person baseline assessment with neuroimaging. Neuroimaging sequences will be used to assess the primary and secondary outcomes. Participants will complete some additional study measures (see below). Caregivers will complete several measures pertaining to their child’s pain and child’s worries in addition to a measure of their own psychological symptoms. To minimize burden, participants and caregivers will be given the option to either complete the majority of measures up to 5 days before this visit using their own electronic device (to reduce the length of the in-person visit; see details below).

Psychometrically validated instruments (self-report unless otherwise indicated) sensitive to change in pediatric health studies will be used. All assessments will be delivered directly to participants and caregivers and collected via autonomous self-report in REDCap, either during the in-person neuroimaging study visits or 1-5 days beforehand. The research coordinator will oversee data collection to ensure data is complete and collected in a timely manner. The study coordinator performing outcome assessments will be blinded to group assignment.

During the neuroimaging portion of the assessment, participants will use a sliding scale to report their current pain intensity, pain unpleasantness, and anxiety. Specifically, participants will enter the MRI scanner, where scans of brain structures and cerebral blood flow will be performed while participants provide current pain and anxiety ratings (∼25 minutes). Participants will then exit the scanner to undergo the WL-SPT, a validated, non-invasive procedure for youth ages 8-16 to create visceral pain sensations which has been previously used by the research team to understand neural mechanisms of FAPD pain [1].

Participants will ingest water until they have achieved complete fullness eliciting discomfort (∼5 minutes). Immediately following the WL-SPT, participants will rate their current pain and anxiety. Then, participants will re-enter the scanner and repeat a similar series of scans (∼20 minutes) with current pain and anxiety ratings. At the completion of all scans, participants will rate their current pain and anxiety once more. Post-assessment measures will follow these same procedures.

#### 2.5.1. Primary Outcomes (collected at pre and post assessments)

Blood oxygenation level dependent (BOLD) resting-state fMRI (rs-fMRI) will be used to assess functional connectivity between the left and right amygdala and brain regions implicated in the FAPD pain experience [1, 15–18] prior to and following WL-SPT, in those undergoing ADAPT versus waitlist.

#### 2.5.2. Secondary Outcomes (collected at pre and post assessments)

Similarly, the potential regional cerebral blood flow (CBF) prior to and following WL-SPT, in the ADAPT group versus waitlist will be assessed with the 3D PASL MRI perfusion technique.

#### 2.5.3. Exploratory Outcomes (collected at pre and post assessments)

We will explore if changes in functional connectivity (BOLD) and regional CBF following ADAPT will correspond to reductions in pain (intensity and unpleasantness) and anxiety ratings. We will also explore the effect of pain on the connectivity of all 17 brain networks as defined by Yeo et al. [24].

##### Magnetic Resonance Imaging (MRI)

MRI data will be acquired on a Siemens Prisma 3T scanner with a 64-channel head/neck coil. Image quality using this scanner has been certified by an MRI physicist (DZ and YZ) based on both human brain and phantom scans. The following three sequences will be used in this study:

1. <u>3D T1-weighted Magnetization-Prepared-Rapid-Acquisition-of-Gradient-Echo (</u>3D MPRAG<u>E) sequence</u> (5 min 12 sec) will be used to visualize anatomical regions and to isolate regions of interest for the analyses of functional connectivity and CBF. Acquisition parameters: voxel size = 1×1×1 mm^3^, field of view (FOV) = 256 × 256 mm^2^, 208 sagittal slices, time of repetition (TR) = 2300 ms, time of echo (TE) = 2.98 ms, time of inversion (TI) = 900 ms, flip angle (FA) = 9°, and parallel imaging acceleration factor = 2. Brain parcellation, regional and whole-brain volume calculation will be performed using FreeSurfer [25]. The accuracy of the procedures will be confirmed by visual inspection.
2. <u>rs-fMRI (12 min 2 sec) will be used to assess functional connectivity</u>. In a dim-light environment, a 12-minute rs-fMRI scan will be acquired while the subjects are asked to relax and have their eyes closed [26]. An eye-closed condition will likely be easier to maintain for our subjects [27, 28]. Acquisition parameters: Isotropic voxel size of 2.5 × 2.5 × 2.5 mm^3^, 60 contiguous axial slices, FOV = 22 cm × 22 cm, TE = 30 ms, TR = 1100 ms, FA = 63°, 4 dummy time points discarded, 650 time points, and simultaneous multislice acceleration factor = 4. Data will be analyzed using AFNI [29]. Slice-timing and motion corrections will be applied. Seed-based functional connectivity to the left/right amygdala will be analyzed based on the correlation between the mean time-course at the left/right amygdala and other voxels in the rest of the brain. Similar seed-based connectivity analyses can be carried to other brain regions.

In addition to examining functional connectivity to the amygdala, we will also calculate the network connectivity of the 17 functional networks defined in the atlas developed by Yeo et al. [24], which is based on rs-fMRI datasets from 1,000 healthy young adults as part of our exploratory analysis. Since pain is a heterogenous experience with multiple networks involved, we will explore these 17 networks broadly and also anticipate specific patterns of change emerging for the default mode network [1] defined in this atlas.

1. <u>3D Pulse Arterial Spin Labeling (</u>3D PASL) sequence at a perfusion mode of FAIR QII (5 min 24 sec) will be used to measure regional CBF [30, 31]. Acquisition parameters: 3D acquisition with FOV = 240 × 240 mm^2^, matrix size = 64 × 64, 32 4.5-mm axial slices, TR = 4000 ms, TE = 20.26 ms, bolus duration = 800 ms, inversion time = 2000 ms,10 control-label pairs of images acquired and averaged to improve signal/noise ratio, and fat saturation. The relative CBF maps will be generated based on the relatively difference of the control and label images.

The 3D MPRAGE images will be collected at the beginning of the scan. To assess the effect of WL-SPT on functional connectivity and CBF, the rs-fMRI and ASL images will be collected before and after the WL-SPT (Fig 4). Participants are instructed not to consume food or water within one hour prior to the MRI scan. Total MRI scanning time is approximately 45 minutes per session to minimize participant burden.

**Fig 4.**
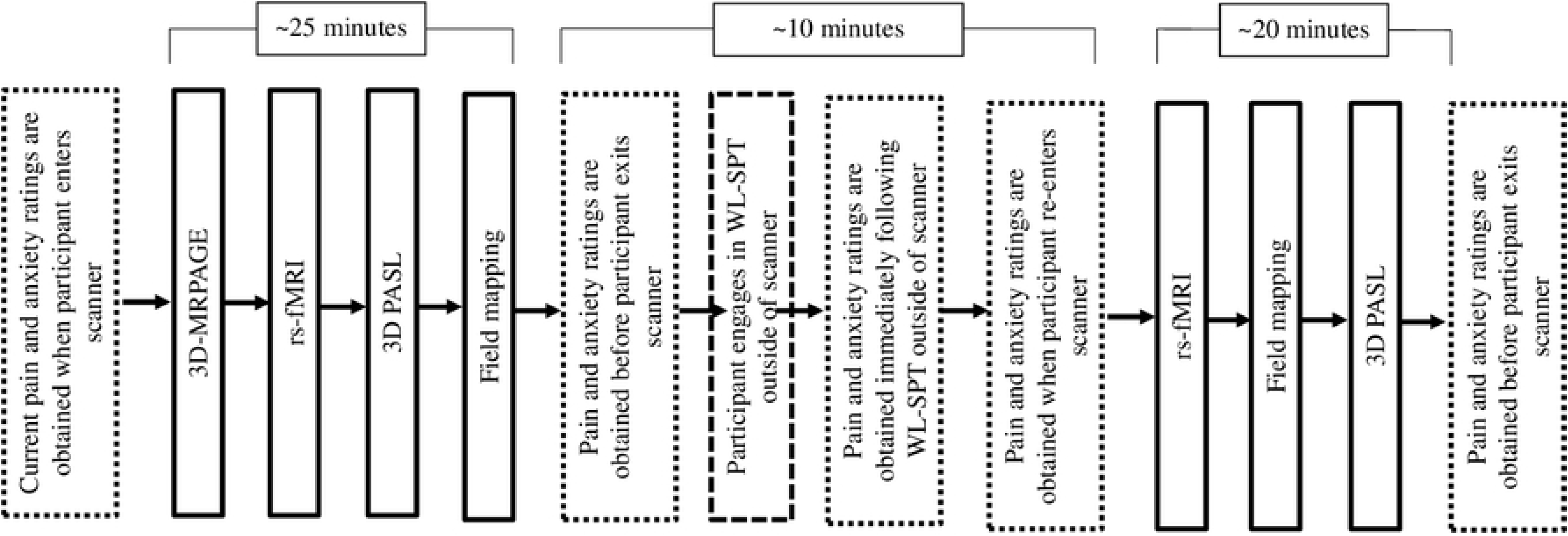
MRI Procedure. The timing scheme of pain and anxiety ratings, MRI and WL-SPT.

#### 2.5.3. Exploratory Outcomes (collected at pre and post assessments)

##### Pain Intensity and Unpleasantness Visual Analog Scales (VAS) [32, 33]

Pain intensity is associated with nociceptive processing and is a common measure of treatment response [34]. Pain unpleasantness is related to affective network activity and is highly responsive to meditation [35]. These will be collected using a physical sliding scale during neuroimaging.

##### State Anxiety (VAS) [36]

0-10 self-report of how anxious the child is feeling in the present moment. This will be collected with a physical sliding scale during neuroimaging.

#### 2.5.4. Other Related Measures (collected at pre and post assessments unless otherwise specified)

##### Fullness Rating Scale [37]

Youth will be asked to indicate how full they felt after water ingestion by selecting from images representing different levels of fullness, from empty (coded 0) to full (coded 4). This will be collected in person during neuroimaging.

##### Edinburgh Handedness Inventory [38]

A validated measure that assesses the dominance of a person’s right or left hand in everyday activities. Given our pediatric sample, one original item was removed (striking a match).

##### Menstruation Query

Female participants who have reached menarche will be asked if they are currently menstruating and if so, how much pain is associated (using a visual sliding scale). During screening, participants will be asked if they have achieved menarche. If participants indicate that they have, we will query the date of their last menstrual cycle.

##### Peterson Pubertal Developmental Scale (PDS) [39]

A valid and reliable pubertal status assessed via clinician interview.

##### Concomitant Medication Form

Concomitant medication information (medication name, reason for taking, unit, frequency, route, etc.) and therapies will be obtained via chart review prior to the baseline visit. Study staff will confirm with caregivers that the information is correct and up to date at the baseline and post assessment visits. Any changes in medication from baseline to post assessment will be documented.

#### 2.5.5. Participant Characteristics and Demographic Factors (collected at pre and post assessments unless otherwise indicated)

##### Child Pain History & Sociodemographic Factors

Demographic factors, school absences, pain duration, location, concomitant psychological treatments, and insurance type. Birth date, sex, gender, race, and ethnicity of participant will also be collected.

##### Primary Care Provider

The name of participant’s primary care provider and the name of their practice, along with contact information will be collected at the pre assessment.

#### 2.5.6. Screening Measures (collected at screening, prior to study enrollment)

##### Rome IV FAPD Diagnosis Checklist [21] (medical provider report)

FAPD criteria based on the Rome IV criteria [21]. FAPD includes irritable bowel syndrome (IBS), functional dyspepsia, abdominal migraine, and FAPD-not otherwise specified. To meet the criteria for FAPD, a child must endorse continuous or episodic pain at least 4 times in a month that does not exclusively occur during a physiological event (e.g., eating, menses) for a period of 2 months or longer, and that cannot be fully explained by another medical condition after appropriate evaluation. This questionnaire will allow us to group our samples into specific diagnostic subtypes of FAPD.

##### Generalized Anxiety Disorders-7, GAD-7 [19]

A seven-item measure assessing for generalized anxiety symptoms in the last two-weeks. The measure has a cut-off score for moderate anxiety ≥ 10. This measure has been validated for use in pediatric populations [40]. This measure is also collected at the post assessment.

##### Patient Health Questionnaire-9 [22]

A validated and reliable measure of depressive symptoms in the past 2 weeks. Categories of severity are as follows: 0-4: Minimal Depression, 5-9: Mild Depression, 10-14: Moderate Depression, 15-19: Moderately Severe Depression, 20-27: Severe Depression. This measure is also collected at the post assessment. Those endorsing thoughts of death on this measure will undergo a comprehensive safety assessment to rule out active suicidal ideation. Youth will be excluded from participating in the study (and referred for additional care) if they report current active suicidal ideation or if their score is 22 or higher.

##### MRI Safety and Screening

Research staff will screen for MRI contraindications to determine if participant can safely complete MRI protocol.

##### Claustrophobia Screening

Research staff will screen for signs of claustrophobia to determine if the participant is claustrophobic. A brief interview to further assess for a fear of enclosed spaces will be administered. This interview was adapted from a portion of the Anxiety Disorder Interview Schedule-Child Report [41], a validated, semi structured diagnostic interview to assess for the presence of child psychological disorders. Responses on this module will help to determine eligibility for the study and will be used in an attempt to reduce instances of unanticipated participant anxiety during the neuroimaging scan. This process has been effective in our pilot neuroimaging research [1] with this population as we did not lose a single participant due to scanner-related anxiety.

#### 2.5.7. Pain Related Measures (collected at pre and post assessments)

##### **Visual Analog Scale (VAS)** [32] **for pain**

Participants will report their average, highest, and lowest pain levels in the past two weeks. If the electronic or physical VAS slider cannot be used (e.g. due technical difficulties), the numeric rating scale (NRS) will be used. Parents will also provide an assessment of their child’s pain in the past two weeks.

##### Other Pain Locations

Participants will be queried about other pain sources besides abdominal pain.

#### 2.5.8. Functional Disability Related Outcomes (collected at pre and post assessments)

##### Functional Disability Inventory (FDI) [42]

A 15-item measure of physical/daily function in last few days. This measure has been validated in pediatric chronic pain [43] and used in pediatric FAPD samples [5, 9]. Child and parent versions will be obtained.

##### NIH PROMIS Pain Related Disability Pain Interference Scale[44]

A validated measure of functional impairment due to pediatric pain in the past 7 days.

#### 2.5.9. Mental Health Related Measures (collected at pre and post assessments)

##### Screen for Child Anxiety Related Disorders, SCARED [45]

Child anxiety (reported by child and parent) in the past 3 months; ≥25 is indicative of clinical anxiety. This measure has been recommended for use by the American Academy of Pediatrics [46] and has been validated in pediatric chronic pain [45, 47, 48] and used in pediatric FAPD samples [5, 9].

##### Child COVID-19 Related Distress

A child-reported item assessing the child’s distress in relation to the COVID-19 pandemic on a sliding scale from 0-10, with anchors labeled “no distress” and “extreme distress”. This measure was added prior to launching the study and following the COVID-19 pandemic.

##### COVID-19 Exposure and Family Impact Survey (CEFIS) [49]

A measure from the Center for Pediatric Traumatic Stress to capture the impact of the COVID-19 pandemic on families and caregivers. This measure was added following the COVID-19 pandemic, which occurred prior to launching the study.

##### Self-Efficacy Pain Scale-Child Version [50]

A validated and reliable 7-item measure of child self-efficacy (reported by both child and parent) when in pain.

##### Affective Reactivity Index (ARI)-Self-Report

A validated 7-item measure of child irritability in the last 7 days. Participants will complete this measure as self-report, and caregivers will provide their report of the child.

##### Pain Catastrophizing Scale for Children [51]

A validated 13-item measure of maladaptive beliefs about pain and feelings experienced when in pain. Participants will complete this measure as self-report, and caregivers will provide their report of the child.

##### Depression Anxiety Stress Scales [52]

A validated and reliable measure with 21 items completed by the caregiver of assessing for self-reported caregiver depression, anxiety, and tension/stress.

##### Adverse Childhood Experiences (ACEs) [53]

A valid, 9-item measure used by the National Survey of Children’s Health that assesses for adverse events occurring in childhood before the age of 18.

#### 2.5.10. Concomitant Care

Any concomitant interventions (i.e., medical, and psychological) experienced by the participants (either allowed (e.g., pharmacological treatments for FAPD) or prohibited (psychological therapy for anxiety or pain), see section *2.2* *Eligibility Criteria*) will also be recorded at baseline and post assessments. Participants will provide the reason and duration of concomitant interventions at the baseline and post assessment visits.

#### 2.5.11. Adverse Event Queries

Participants will be queried about any experienced adverse events during their second session of ADAPT (if randomized to ADAPT treatment group). This form will also be administered at post assessment. For those randomized to waitlist, they will complete this form at post assessment, and then at ADAPT sessions 2 and 6 (if they opt to complete ADAPT).

##### Adverse Event Query Form

A form to be completed via clinician or research coordinator interview assessing for adverse events experienced by participants.

### 2.6. Intervention

#### 2.6.1. The Aim to Decrease Anxiety and Pain Treatment (ADAPT)

ADAPT is a psychological intervention that, in the current investigation, integrates mindfulness meditation with cognitive behavioral therapy approaches for managing pain and anxiety to improve patient outcomes (see Table 1). ADAPT was originally developed from a CBT protocol for pain management [12] and from the “Cool Kids” program for anxiety [11]. The current iteration integrates mindfulness meditation strategies from an established protocol [13]. ADAPT is primarily a child-focused, individual therapy with some caregiver involvement. The duration of ADAPT is 6 six weeks, although up to eight weeks will be permitted to account for scheduling conflicts and participant illness. In the current study, there will be two live videoconference (Zoom) sessions with an interventionist (60 minutes) once per week for the first two weeks, followed by 4 weeks of self-paced web modules (30 minutes per week) in conjunction with interventionist support (15-30 minutes per week). Prior to the study launch (as a result of the COVID-19 pandemic), ADAPT was revised to be delivered fully remotely (e.g., live, in-person sessions to be delivered via videoconferencing). In the event of technical difficulties, live sessions can also be completed over phone calls. Preliminary testing has shown that ADAPT is feasible and successfully reduces pain-related disability and anxiety over time [9, 14]. The interaction with the interventionist has been shown to be a key component engaging youth in the program [54], and ADAPT has shown to be associated with improvements in caregiver anxiety [55]. All participants will receive or be offered ADAPT, either immediately following baseline assessment and randomization, or after completing the post assessment if participants are randomized to waitlist.

**Table 1.**
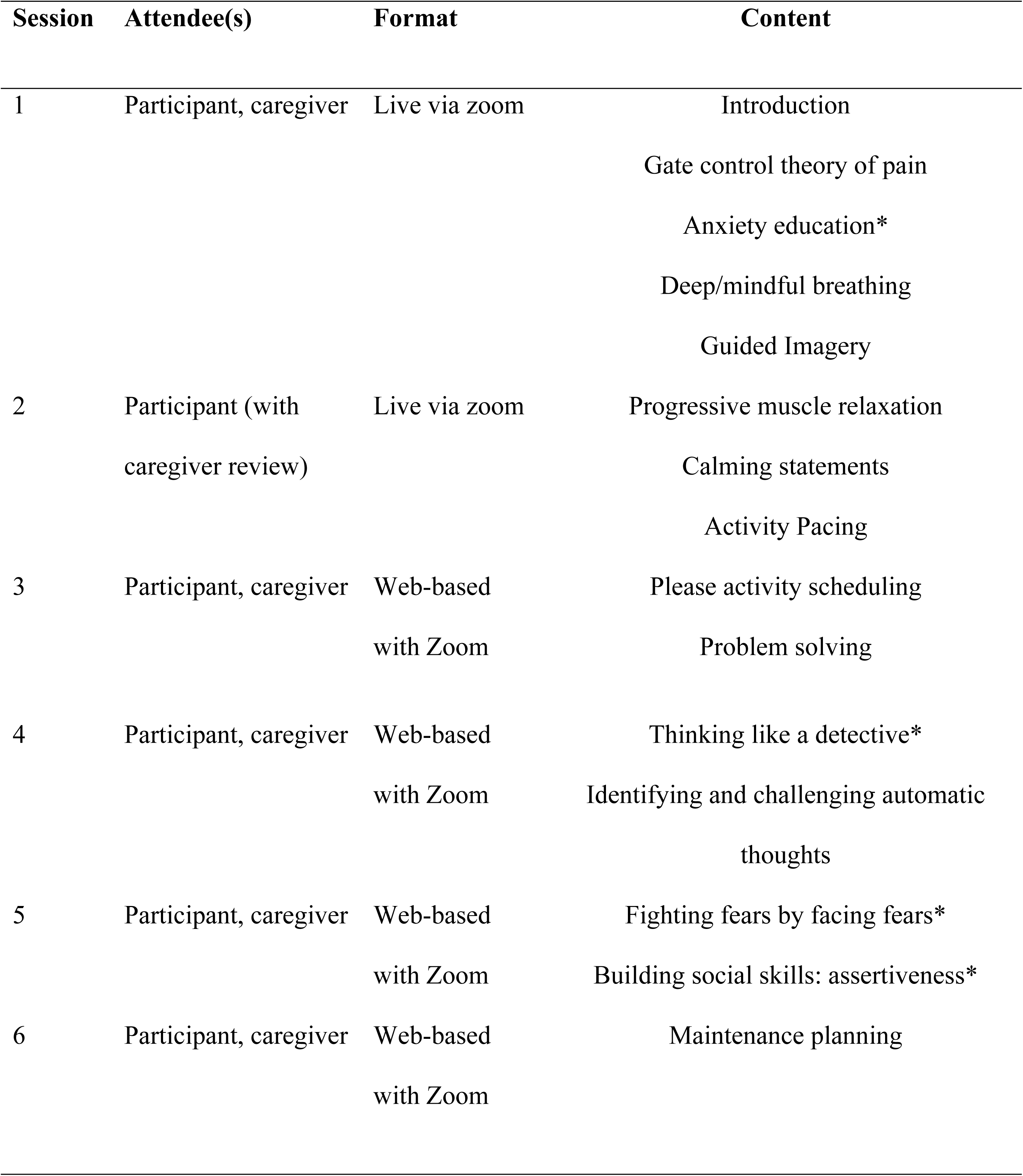
ADAPT Protocol. Note. Live sessions occur via HIPPA-compliant Zoom videoconferencing (1 hour). Web-based sessions with zoom are self-paced modules (approximately 30 minutes) and therapist support (approximately 15-30 minutes). All participants are permitted to continue their medical care as usual throughout the duration of the study. *Drawn from anxiety-specific treatment protocol, “Cool Kids” [11]. The remaining content is adapted from a pain coping skills program [12] or a mindfulness meditation protocol [13].

#### 2.6.2 Treatment integrity

All sessions will be video recorded (with patient permission) and stored. Treatment integrity will be assessed using video recordings of videoconference sessions by independent evaluators (e.g., doctoral students or medical trainees). Integrity scores (0–100%) used in prior research [12, 14] will be obtained. Items assessed included delivery of session-specific skills and assessment of participant comprehension. There will also be regular supervision of the interventionist with a licensed psychology to review protocol and insure treatment fidelity.

#### 2.6.3 Waitlist group

Participants in the waitlist arm will follow their gastroenterologists’ recommendations for FAPD management. Waitlist participants will not be permitted to receive psychological care for pain or anxiety outside of the study during their participation in order to minimize possible similar treatment effect on waitlist control group, and will be offered ADAPT following their post assessment as a courtesy.

### 2.7. Data Management and Monitoring

An external data monitoring committee comprising of 3 external MSU faculty with specialties in bio-behavioral statistics, global neuropsychiatry, and neuroscience will meet quarterly to assess study progress. The NCCIH will provide oversight via yearly study site visits through Westat.

To ensure confidentiality, identification numbers will be used in lieu of names and data will be kept in locked files in the PI’s research location or on a secure computer that is designated specifically for the purposes of this project. All data (except for MRI data) will be saved into REDCap, a password-protected database. To assure the accuracy of data entry, data will be verified in real-time by study staff via database-incorporated data entry checks. Data output will be stored on a network devoted solely to the research activities associated with MSU’s Biomedical Research Informatics Core (BRIC). Electronic data stored on MSU’s network is backed up nightly by BRIC. Only the PI and designated members of her team will have access to the final trial dataset. We expect to publish the results and share them on the ClinicalTrials.gov database.

The High-Performance Computing Center (HPCC) will be used for checking, storing, and analyzing MRI data. The HPCC is maintained by MSU’s Institute for Cyber-Enabled Research (ICER). The MRI data will also be stored on a secure server, backed up nightly, and will only be accessible to study staff.

While all efforts will be made to minimize missing data through the use of electronic data capture and real-time adherence data collection, missing data is still inevitable in RCTs. Thus, we will employ several strategies to handle missing data with specific attention on how to handle missing not-at-random data. Specifically, missing data will be handled via ML estimation with auxiliary correlate inclusion [56, 57]. Missing not at random will be addressed with mixture MNAR methods [58–60].

### 2.8 Statistical Analysis Plan

Primary and secondary outcome measures are obtained from neuroimaging data. The brain regions selected have been shown in prior literature to be related to chronic pain [1, 61] and respond to psychological interventions for the management of chronic pain conditions [62].

The 3D T1-weighted MPRAGE images will be segmented via FreeSurfer “recon-all” [25]. The segmentation will be used in rs-fMRI functional connectivity and CBF analyses.

For rs-fMRI data analysis, image pre-processing and data analysis will be accomplished by Analysis of Functional Neuroimages (AFNI) [63] and FreeSurfer [25] softwares. Signal time course spikes will be detected and removed. Data points with excessive motion will be identified (normalized motion derivative > 0.5 or vox outliers > 10%) and modeled as regressors in subsequent processing. Acquisition timing differences across slice locations will be corrected.

Rigid-body motion correction will be applied in three translational and three rotational directions. Translational and rotational estimates, as well as their derivatives, will be modeled as regressors for the subsequent noise regression step. For each subject, spatial blurring with a full-width half-maximum of 4mm will be used to reduce random noise. CSF and white matter mean signals will be modeled as nuisance variables. A band-pass filter (0.009 Hz-0.08 Hz) will be applied. For the time course at each voxel location, temporal noise discussed above will be removed via “3dDeconvolve” feature in AFNI [63].

**Functional Connectivity Analysis:** Each functional connectivity map will be warped to the standard MNI152 space via AFNI. Correlation coefficients for each subject will be transformed using Fisher’s Z. In the overall group analysis on the effect of WL-SPT, paired t-tests between the pre and post WL-SPT functional connectivity maps will be conducted. ANOVAs will be conducted on the functional connectivity differences between the ADAPT completers and those in the waitlist condition. Statistically significant regions will be identified based on p<0.05 after multiple comparison correction, which will be estimated according to Monte Carlo simulations in AFNI [63]. Changes between the amygdala and other network regions (e.g., thalamus) that are implicated in pain processing are expected following ADAPT versus waitlist (both following WL-SPT, and while at rest).

**Regional CBF analyses:** The relative CBF maps will be generated for each PASL scan. We will use a whole-brain search to identify areas associated with connectivity with the amygdala. The CBF maps will be aligned to the T1 MPRAGE. The mean CBF values on the regions segmented by FreeSurfer (Fischl, 2012) will be calculated. The percentage change at each region due to WL-SPT will be calculated. ANOVAs testing the percentage change of CBF at the brain regions will be completed to compare the ADAPT completers to those in the waitlist condition.

Pain-related brain CBF is expected to diminish for those that complete ADAPT compared to the waitlist condition (Aim 2) following the WL-SPT and while at rest.

**Exploratory Analyses:** Neural mechanisms associated with a positive treatment response (decreased pain intensity, unpleasantness, and state anxiety (after the WL-SPT and while at rest) after ADAPT will be identified. Normalization of abnormal connectivity patterns are predicted to categorize changes in participant-reported pain and anxiety. Specifically, decreases in brain CBF and reductions in functional connectivity will be examined in relation to improvements in pain and anxiety post treatment using multiple (linear) regression.

Given that other factors such as pubertal status and subjective pain levels could influence the outcome, these factors will be included in the analysis as control covariates. Each participant’s exact age will be recorded at assessment allowing personnel to control for age as an additional covariate should the need become apparent during preliminary analyses.

Network-based exploratory connectivity analysis will also be conducted. The cortical nodes of the 17 networks identified by Yeo and colleagues (2011) will be identified via Freesurfer segmentation (Fischl, 2012). Correlation between each pair of nodes in each network will be calculated in MATLAB [64]. The connectivity within each network is the average of the correlations of all pairs of nodes within a network. In assessing whole-brain connectivity, the global average of the absolute r values between all nodes will be taken (to account for networks known to be anticorrelated).

All correlation coefficients of each subject will be transformed using Fisher’s Z using SPSS (IBM SPSS Statistics for Windows, Version 26.0). In the overall group analysis on the effect of WL-SPT, paired t-tests between the pre and post WL-SPT functional connectivity of each network and global connectivity will be conducted. ANOVAs will be conducted to investigate functional connectivity differences between the ADAPT completers and those in the waitlist condition.

#### 2.8.1. Sample size and randomization specifications

We used the following tool (http://neuropowertools.org/neuropower/neuropowerinput/) to conduct sample size calculations for the rs-fMRI portion of the study. Power calculations were based on prior studies by research members on functional connectivity data 1) pre and post psychological therapy for pediatric pain (migraine) [65], and 2) comparing individuals with pediatric pain (migraine) to healthy controls [66] .

For within group changes, we relied on the pre/post data following a psychological therapy for pediatric pain (migraine) [65]. A total sample size of 34 is required for power of .8 and p < 0.05 to observe at least moderate (e.g. mean effect size difference of 0.4 or greater) effects. For between group changes, we utilized data comparing youth with chronic pain (migraine) to healthy controls [66]. Based on these data, a total of 35 subjects are required for power of .8 and p < 0.05.

### 2.9 Interim analyses and stopping rules

There are no planned interim analyses. Dr. Barber Garcia will assess adherence and retention. If the treatment is deemed to be ineffective, unsafe, futile, or if there is evidence of poor study performance (e.g., slow accrual, high losses-to-follow-up, and poor quality control), a safety review may be warranted. This action would occur in consultation with the study mentors (Zhu, Arnetz), the IMC, and NCCIH.

## Discussion

FAPD are a set of common and chronic abdominal pain conditions [20, 21, 67] generally categorized by hyperconnectivity between the amygdala and other regions of the brain implicated in cognition and pain-processing [1, 61, 68, 69]. Further, FAPD are associated with significant psychosocial impairment and may persist for a subset of youth [70, 71]. Pain-related outcomes of FAPD are worse in the presence of comorbid anxiety [5, 72]. Further, anxiety, in combination with clinically elevated levels of pain and pain-related disability, are associated with persistent functional impairment [8]. Anxiety also diminishes impact of pain-focused CBT [9], which is among the most widely researched approach to care for pediatric chronic pain conditions [73]. Fortunately, a coping skills program called the Aim to Decrease Anxiety and Pain Treatment (ADAPT) has been developed [10] that addresses comorbid anxiety and chronic pain symptoms for youth with FAPD. The program consists of a blend of CBT strategies and was infused with mindfulness meditation to be delivered by a trained interventionist weekly for a period of six weeks [10, 14]. ADAPT has shown to be superior to improving pain-related disability and anxiety as compared to standard care in a randomized clinical trial [14]. However, the potential neural mechanisms underlying the effect of this treatment are unknown. Once such factors are investigated through our neuromechanistic trial, we will have an enhanced understanding of mechanisms underlying treatment response to psychological interventions addressing pain and comorbid anxiety.

Our trial utilizes a validated visceral symptom provocation task, the water loading symptom provocation task (WL-SPT) [37]. We have intentionally used this task during neuroimaging the brains of youth with FAPD [1], who otherwise may or may not be experiencing an active pain flare at the time of imaging. The WL-SPT is known to induce sensations including pain intensity and pain unpleasantness similar to, but less severe than, an active pain flare [37]. Therefore, in our current neuromechanistic trial, we will compare youth functional connectivity between the amygdala and other regions of the brain after ADAPT versus waitlist during after the WL-SPT, in addition to rest. To enhance the rigor of the results, visual analog measures of pain severity/unpleasantness [32, 33] and anxiety [36] will be obtained throughout the scanning. Such a research design employing a validated visceral symptom provocation task has the potential to advance the literature.

We expect minimal risk and potential benefits to participants and their families following receipt of the ADAPT program, which is an evidence-based program to improve pain-related disability and anxiety in youth with FAPD [14]. Limitations of the current study design include lack of active treatment comparator and a relatively modest sample size. We also recognize functional connectivity is based on surrogates of neural activity (i.e., changes in blood flow in response to neural activity); thus, findings should be interpreted within that lens. Future planned research includes dissemination and implementation trials, examining sustainability of treatment effects and associated neural mechanisms, and investigating neural mechanisms underlying components of treatment response, similar to work currently underway in pediatric migraine [65].

Given that FAPD are common [2] and costly [74] conditions associated with changes in neural processing [1], and effective programs such as ADAPT are available [14], it is important to understanding neural mechanisms of treatment response. This is particularly true in the adolescent population as FAPD [5, 8] and anxiety [75] increases and can become more severe as children age and transition to adolescence. This is a period marked by pubertal and developmental changes that may alter changes in neural functioning associated with pain processing [76]. Therefore, additional investigation of the impact of pubertal status on pain processing following effective treatments will be important in future research. Though this work is specific to FAPD, findings may be a guide to enhance the understanding of treatment response of other pediatric chronic pain conditions (which are also generally categorized by enhanced functional connectivity between the amygdala and other regions associated with pain) [77, 78]. This is critically important as chronic pain conditions are the most common childhood chronic illnesses and most expensive health problem facing children in the US [79].

## Conclusion

In conclusion, this is the first neuromechanistic RCT of a tailored psychological treatment to target pain and anxiety in youth with FAPD. Results aim to improve understanding of mechanisms underlying response to psychological treatments for FAPD.

## Data Availability

No datasets were generated or analysed during the current study. All relevant data from this study will be made available upon study completion.

## Acknowledgements

We thank the gastroenterology team at Helen DeVos Children’s Hospital including Drs. Vanessa Cardenas-Soto, Peter Freswick, Deborah Cloney, Sarah Henkel, Amie Hinshaw, nurse practitioner Ashley Verhey, and social workers Mackenzie Mullins and Jamie Bart for supporting the recruitment efforts and execution of the study. We thank Drs. Amy Pugh, Alexander Redman, Christina Varghese, and Leo Fawaz for providing on-call services. We thank Mallet Reid and Sukhesh Sudan for their assistance with randomization. We thank Emme Bourassa, Kara Nearanz, and Caitlin Wilterink for their support as Corewell Health research assistants. We thank Elise Cheney-Makens for her support with recruitment. We thank Robert Jones, Stacy Clegg, Becky Ruiter, Carrie Popilek, Michele Harvey, Steven Zomberg, and Crista Bradley for their support as MRI technologists on the project.

## Notes

### Competing Interest Statement

The authors have declared no competing interest.

### Clinical Trial

NCT03518216

### Clinical Protocols

https://classic.clinicaltrials.gov/ProvidedDocs/16/NCT03518216/Prot_SAP_000.pdf

### Author Declarations

This study is approved by Michigan State University’s institutional review board, with Helen DeVos Children’s Hospital/Corewell Health relying on the primary IRB.

## References

1. Cunningham NR, Nahman-Averbuch H, Lee GR, King CD, Coghill RC. Amygdalar functional connectivity during resting and evoked pain in youth with functional abdominal pain disorders. J Pain. 2022;163(10):2031–43.

2. King S, Chambers CT, Huguet A, MacNevin RC, McGrath PJ, Parker L, et al. The epidemiology of chronic pain in children and adolescents revisited: a systematic review. J Pain. 2011;152(12):2729–38.

3. Walker LS, Sherman AL, Bruehl S, Garber J, Smith CA. Functional abdominal pain patient subtypes in childhood predict functional gastrointestinal disorders with chronic pain and psychiatric comorbidities in adolescence and adulthood. J Pain. 2012;153(9):1798–806.

4. Størdal K, Nygaard EA, Bentsen BS. Recurrent abdominal pain: a five-year follow-up study. Acta Paediatr. 2005;94(2):234–6.

5. Cunningham NR, Cohen MB, Farrell MK, Mezoff AG, Lynch-Jordan A, Kashikar-Zuck S. Concordant parent-child reports of anxiety predict impairment in youth with functional abdominal pain. J Pediatr Gastroenterol Nutr. 2014;60(3):312–7.

6. Dorn LD, Campo JC, Thato S, Dahl RE, Lewin D, Chandra R, et al. Psychological comorbidity and stress reactivity in children and adolescents with recurrent abdominal pain and anxiety disorders. J Am Acad Child Adolesc Psychiatry. 2003;42(1):66–75.

7. Dufton LM, Dunn MJ, Compas BE. Anxiety and somatic complaints in children with recurrent abdominal pain and anxiety disorders. J Pediatr Psychol. 2009;34(2):176–86.

8. Cunningham NR, Jagpal A, Peugh J, Cohen MB, Farrell MK, Mezoff AG, et al. Using a simple risk categorization to predict outcomes in pediatric functional abdominal pain at 6 months J Pediatr Gastroenterol Nutr. 2017;64(5):685–90.

9. Cunningham NR, Jagpal A, Tran ST, Kashikar-Zuck S, Goldschneider KR, Coghill RC, et al. Anxiety Adversely Impacts Response to Cognitive Behavioral Therapy in Children with Chronic Pain. J Pediatr. 2016;171:227–33.

10. Cunningham NR, Nelson S, Jagpal A, Moorman E, Farrell M, Pentiuk S, et al. Development of the Aim to Decrease Anxiety and Pain Treatment for Pediatric Functional Abdominal Pain Disorders. J Pediatr Gastroenterol Nutr. 2018;66(1):16–20.

11. Lyneham HJ, Abbott MJ, Wignall A, Rapee RA. The Cool Kids Anxiety Treatment Program. 2003.

12. Kashikar-Zuck S, Ting TV, Arnold LM, Bean J, Powers SW, Graham TB, et al. Cognitive behavioral therapy for the treatment of juvenile fibromyalgia: a multisite, single-blind, randomized, controlled clinical trial. Arthritis Rheum. 2012;64(1):297–305.

13. Broderick P. Learning to breathe. Oakland, CA: New Harbinger Publications; 2013.

14. Cunningham NR, Kalomiris A, Peugh J, Farrell M, Pentiuk S, Mallon D, et al. Cognitive Behavior Therapy Tailored to Anxiety Symptoms Improves Pediatric Functional Abdominal Pain Outcomes: A Randomized Clinical Trial. J Pediatr. 2021;230:62–70.e3.

15. Raichle ME, MacLeod AM, Snyder AZ, Powers WJ, Gusnard DA, Shulman GL. A default mode of brain function. Proceedings of the national academy of sciences. 2001;98(2):676–82.

16. Baliki MN, Chialvo DR, Geha PY, Levy RM, Harden RN, Parrish TB, et al. Chronic pain and the emotional brain: specific brain activity associated with spontaneous fluctuations of intensity of chronic back pain. J Neurosci. 2006;26(47):12165–73.

17. Baliki MN, Geha PY, Apkarian AV, Chialvo DR. Beyond feeling: chronic pain hurts the brain, disrupting the default-mode network dynamics. J Neurosci. 2008;28(6):1398–403.

18. Cunningham NR, Kashikar-Zuck S, Coghill RC. Brain mechanisms impacted by psychological therapies for pain: identifying targets for optimization of treatment effects. PAIN Reports. 2019;4(4):e767.

19. Spitzer RL, Kroenke K, Williams JB, Löwe B. A brief measure for assessing generalized anxiety disorder: the GAD-7. Arch Intern Med. 2006;166(10):1092–7.

20. Walker LS, Lipani TA, Greene JW, Caines K, Stutts J, Polk DB, et al. Recurrent abdominal pain: symptom subtypes based on the Rome II Criteria for pediatric functional gastrointestinal disorders. J Pediatr Gastroenterol Nutr. 2004;38(2):187–91.

21. Koppen IJ, Nurko S, Saps M, Di Lorenzo C, Benninga MA. The pediatric Rome IV criteria: what’s new? Expert Rev Gastroenterol Hepatol. 2017;11(3):193–201.

22. Kroenke K, Spitzer RL, Williams JB. The PHQ-9: validity of a brief depression severity measure. J Gen Intern Med. 2001;16(9):606–13.

23. Kroenke K, Spitzer RL, Williams JB, Löwe B. An ultra-brief screening scale for anxiety and depression: the PHQ-4. Psychosomatics. 2009;50(6):613–21.

24. Yeo BT, Krienen FM, Sepulcre J, Sabuncu MR, Lashkari D, Hollinshead M, et al. The organization of the human cerebral cortex estimated by intrinsic functional connectivity. J Neurophysiol. 2011;106(3):1125–65.

25. Fischl B. FreeSurfer. Neuroimage. 2012;62(2):774–81.

26. Birn RM, Molloy EK, Patriat R, Parker T, Meier TB, Kirk GR, et al. The effect of scan length on the reliability of resting-state fMRI connectivity estimates. Neuroimage. 2013;83:550–8.

27. Fernandez Z, Scheel N, Baker JH, Zhu DC. Functional connectivity of cortical resting-state networks is differentially affected by rest conditions. Brain Res. 2022;1796:148081.

28. Chang SE, Zhu DC. Neural network connectivity differences in children who stutter. Brain. 2013;136(Pt 12):3709–26.

29. Cox RW. AFNI: what a long strange trip it’s been. Neuroimage. 2012;62(2):743–7.

30. Kim SG. Quantification of relative cerebral blood flow change by flow-sensitive alternating inversion recovery (FAIR) technique: application to functional mapping. Magn Reson Med. 1995;34(3):293–301.

31. Wong EC, Buxton RB, Frank LR. Quantitative imaging of perfusion using a single subtraction (QUIPSS and QUIPSS II). Magn Reson Med. 1998;39(5):702–8.

32. McGrath PJ, Walco GA, Turk DC, Dworkin RH, Brown MT, Davidson K, et al. Core outcome domains and measures for pediatric acute and chronic/recurrent pain clinical trials: PedIMMPACT recommendations. J Pain. 2008;9(9):771–83.

33. Price DD, Bush FM, Long S, Harkins SW. A comparison of pain measurement characteristics of mechanical visual analogue and simple numerical rating scales. J Pain. 1994;56(2):217–26.

34. Palermo TM, Eccleston C, Lewandowski AS, Williams ACdC, Morley S. Randomized controlled trials of psychological therapies for management of chronic pain in children and adolescents: an updated meta-analytic review. J Pain. 2010;148(3):387–97.

35. Zeidan F, Martucci KT, Kraft RA, Gordon NS, McHaffie JG, Coghill RC. Brain mechanisms supporting the modulation of pain by mindfulness meditation. J Neurosci. 2011;31(14):5540–8.

36. Kindler CH, Harms C, Amsler F, Ihde-Scholl T, Scheidegger D. The visual analog scale allows effective measurement of preoperative anxiety and detection of patients’ anesthetic concerns. Anesth Analg. 2000;90(3):706–12.

37. Walker LS, Williams SE, Smith CA, Garber J, Van Slyke DA, Lipani T, et al. Validation of a symptom provocation test for laboratory studies of abdominal pain and discomfort in children and adolescents. J Pediatr Psychol. 2006;31(7):703–13.

38. Oldfield RC. The assessment and analysis of handedness: the Edinburgh inventory. Neuropsychologia. 1971;9(1):97–113.

39. Petersen AC, Crockett L, Richards M, Boxer A. A self-report measure of pubertal status: Reliability, validity, and initial norms. J Youth Adolesc. 1988;17(2):117–33.

40. Mossman SA, Luft MJ, Schroeder HK, Varney ST, Fleck DE, Barzman DH, et al. The Generalized Anxiety Disorder 7-item scale in adolescents with generalized anxiety disorder: Signal detection and validation. Ann Clin Psychiatry. 2017;29(4):227–34a.

41. Albano A, Silverman W. The Anxiety Disorders Interview Schedule for Children for DSM-IV: Clinician manual (child and parent versions). San Antonio, TX: Psychological Corporation. 1996;183:649-56.

42. Walker LS, Greene JW. The functional disability inventory: measuring a neglected dimension of child health status. J Pediatr Psychol. 1991;16(1):39–58.

43. Kashikar-Zuck S, Flowers S, Claar R, Guite J, Logan D, Lynch-Jordan A, et al. Clinical utility and validity of the Functional Disability Inventory among a multicenter sample of youth with chronic pain. J Pain. 2011;152(7):1600–7.

44. Varni JW, Stucky BD, Thissen D, DeWitt EM, Irwin DE, Lai J-S, et al. PROMIS Pediatric Pain Interference Scale: an item response theory analysis of the pediatric pain item bank. J Pain. 2010;11(11):1109–19.

45. Birmaher B, Khetarpal S, Brent D, Cully M, Balach L, Kaufman J, et al. The Screen for Child Anxiety Related Emotional Disorders (SCARED): Scale construction and psychometric characteristics. J Am Acad Child Adolesc Psychiatry. 1997;36(4):545–53.

46. Mental Health Screening and Assessment Tools for Primary Care. 2010. In: Addressing Mental Health Concerns in Primary Care [Internet]. Am Acad Pediatr

47. Birmaher B, Brent D, Chiappetta L, Bridge J, Monga S, Baugher M. Psychometric properties of the Screen for Anxiety Related Emotional Disorders (SCARED): A replication study. J Am Acad Child Adolesc Psychiatry. 1999;38(10):1230–6.

48. Mano KEJ, Evans JR, Tran ST, Khan KA, Weisman SJ, Hainsworth KR. The psychometric properties of the Screen for Child Anxiety Related Emotional Disorders in pediatric chronic pain. J Pediatr Psychol. 2012;37(9):999–1011.

49. COVID-19 Exposure and Family Impact Survey (CEFIS). 2020.

50. Bursch B, Tsao JC, Meldrum M, Zeltzer LK. Preliminary validation of a self-efficacy scale for child functioning despite chronic pain (child and parent versions). J Pain. 2006;125(1):35–42.

51. Crombez G, Bijttebier P, Eccleston C, Mascagni T, Mertens G, Goubert L, et al. The child version of the pain catastrophizing scale (PCS-C): a preliminary validation. J Pain. 2003;104(3):639–46.

52. Henry JD, Crawford JR. The short-form version of the Depression Anxiety Stress Scales (DASS-21): Construct validity and normative data in a large non-clinical sample. Br J Clin Psychol. 2005;44(2):227–39.

53. Bethell CD, Carle A, Hudziak J, Gombojav N, Powers K, Wade R, et al. Methods to assess adverse childhood experiences of children and families: toward approaches to promote child well-being in policy and practice. Acad Pediatr. 2017;17(7):S51–S69.

54. Miller AK, Ely SL, Barber Garcia BN, Richardson P, Cunningham NR. Engagement during a Mixed In-Person and Remotely Delivered Psychological Intervention for Youth with Functional Abdominal Pain Disorders and Anxiety. Children (Basel). 2021;8(9).

55. Kalomiris AE, Ely SL, Love SC, Mara CA, Cunningham NR. Child-Focused Cognitive Behavioral Therapy for Pediatric Abdominal Pain Disorders Reduces Caregiver Anxiety in Randomized Clinical Trial. J Pain. 2022;23(5):810–21.

56. Graham JW. Adding Missing-Data-Relevant Variables to FIML-Based Structural Equation Models. Struct Equ Modeling. 2003;10(1):80–100.

57. Enders C. Applied Missing Data Analysis. The Guilford Press 2010.

58. Gottfredson NC, Bauer DJ, Baldwin SA. Modeling Change in the Presence of Non-Randomly Missing Data: Evaluating A Shared Parameter Mixture Model. Struct Equ Modeling. 2014;21(2):196–209.

59. Muthén B, Asparouhov T, Hunter AM, Leuchter AF. Growth modeling with nonignorable dropout: alternative analyses of the STAR*D antidepressant trial. Psychol Methods. 2011;16(1):17–33.

60. Sterba SK, Gottfredson NC. Diagnosing global case influence on MAR versus MNAR model comparisons. Struct Equ Modeling. 2015;22(2):294–307.

61. Simons LE, Moulton EA, Linnman C, Carpino E, Becerra L, Borsook D. The human amygdala and pain: evidence from neuroimaging. Hum Brain Mapp. 2014;35(2):527–38.

62. Simons LE, Pielech M, Erpelding N, Linnman C, Moulton E, Sava S, et al. The responsive amygdala: treatment-induced alterations in functional connectivity in pediatric complex regional pain syndrome. J Pain. 2014;155(9):1727–42.

63. Cox RW. AFNI: software for analysis and visualization of functional magnetic resonance neuroimages. Comput Biomed Res. 1996;29(3):162–73.

64. MATLAB 9.13.0 (R2022b) ed. Natick, Massachusetts: The MathWorks Inc.; 2022.

65. Nahman-Averbuch H, Schneider VJ, 2nd, Chamberlin LA, Kroon Van Diest AM, Peugh JL, Lee GR, et al. Alterations in Brain Function After Cognitive Behavioral Therapy for Migraine in Children and Adolescents. Headache. 2020;60(6):1165–82.

66. Nahman-Averbuch H, Leon E, Hunter BM, Ding L, Hershey AD, Powers SW, et al. Increased pain sensitivity but normal pain modulation in adolescents with migraine. J Pain. 2019;160(5):1019–28.

67. Walker LS, Lipani TA, Greene JW, Caines K, Stutts J, Polk DB, et al. Recurrent abdominal pain: symptom subtypes based on the Rome II criteria for pediatric functional gastrointestinal disorders. J Pediatr Gastroenterol Nutr. 2004;38(2):187–91.

68. Tillisch K, Mayer EA, Labus JS. Quantitative meta-analysis identifies brain regions activated during rectal distension in irritable bowel syndrome. J Gastroenterol. 2011;140(1):91–100.

69. Liu X, Silverman A, Kern M, Ward BD, Li SJ, Shaker R, et al. Excessive coupling of the salience network with intrinsic neurocognitive brain networks during rectal distension in adolescents with irritable bowel syndrome: a preliminary report. Neurogastroenterol Motil. 2016;28(1):43–53.

70. Walker LS, Guite JW, Duke M, Barnard JA, Greene JW. Recurrent abdominal pain: A potential precursor of irritable bowel syndrome in adolescents and young adults. J Pediatr. 1998;132(6):1010–5.

71. Ramchandani PG, Fazel M, Stein A, Wiles N, Hotopf M. The impact of recurrent abdominal pain: predictors of outcome in a large population cohort. Acta Paediatr. 2007;96(5):697–701.

72. Stanford EA, Chambers CT, Biesanz JC, Chen E. The frequency, trajectories and predictors of adolescent recurrent pain: A population-based approach. J Pain. 2008;138(1):11–21.

73. Fisher E, Villanueva G, Henschke N, Nevitt SJ, Zempsky W, Probyn K, et al. Efficacy and safety of pharmacological, physical, and psychological interventions for the management of chronic pain in children: a WHO systematic review and meta-analysis. J Pain. 2022;163(1):e1–e19.

74. Dhroove G, Chogle A, Saps M. A Million-dollar Work-up for Abdominal Pain: Is It Worth It? J Pediatr Gastroenterol Nutr. 2010;51(5):579–83 10.1097/MPG.0b013e3181de0639.

75. APA. Diagnostic and statistical manual of mental disorders (5th ed). Arlington, VA: Am Psychiatr Publ; 2013.

76. Nahman-Averbuch H, Li R, Boerner KE, Lewis C, Garwood S, Palermo TM, et al. Alterations in pain during adolescence and puberty. Trends Neurosci. 2023;46(4):307–17.

77. Simons LE, Moulton EA, Linnman C, Carpino E, Becerra L, Borsook D. The human amygdala and pain: evidence from neuroimaging. Hum Brain Mapp. 2014;35(2):527–38.

78. Erpelding N, Simons L, Lebel A, Serrano P, Pielech M, Prabhu S, et al. Rapid treatment-induced brain changes in pediatric CRPS. Brain Struct Funct. 2016;221(2):1095–111.

79. Groenewald CB, Essner BS, Wright D, Fesinmeyer MD, Palermo TM. The Economic Costs of Chronic Pain Among a Cohort of Treatment-Seeking Adolescents in the United States. J Pain. 2014;15(9):925–33.

